# Ethnic inequalities in multiple long-term health conditions in the United Kingdom: a systematic review and narrative synthesis

**DOI:** 10.1101/2022.03.31.22273224

**Authors:** Brenda Hayanga, Mai Stafford, Laia Bécares

## Abstract

Indicative evidence suggests that minoritised ethnic groups have higher risk of developing multiple long-term conditions (MLTCs), and do so earlier than the white majority population. While there is evidence on ethnicity and single conditions and comorbidities, no review has attempted to look across these from a MLTCs perspective. As such, we currently have an incomplete understanding of the extent of ethnic inequalities in the prevalence of MLTCs. In this systematic review we aimed to 1) describe the literature that provides evidence of ethnicity and prevalence of MLTCs amongst people living in the UK, and 2) summarise the prevalence estimates of MLTCs across ethnic groups. We registered the protocol on PROSPERO (CRD42020218061). Between October and December 2020, we searched ASSIA, Cochrane Library, EMBASE, MEDLINE, PsycINFO, PubMed, ScienceDirect, Scopus, Web of Science, OpenGrey, and reference lists of key studies/reviews. The main outcome was prevalence estimates for MLTCs for at least one minoritised ethnic group, compared to the majority white population. We included studies conducted in the UK reporting on ethnicity and prevalence of MLTCs. To summarise the prevalence estimates of MLTCs across ethnic groups we included only studies of MLTCs that provided estimates adjusted at least for age. Two reviewers screened and extracted data from a random sample of studies (10%). Data were synthesised using narrative synthesis. Of the 7949 studies identified, 84 met criteria for inclusion. Of these, seven contributed to the evidence of ethnic inequalities in MLTCs. Five of the seven studies point to higher prevalence of MLTCs in at least one minoritised ethnic group compared to their white counterparts. Because the number/types of health conditions varied between studies and some ethnic populations were aggregated, the findings may not accurately reflect the true level of inequality. Thus, our conclusions can only be tentative. Future research should consider key explanatory factors, including those at the macrolevel (e.g. racism, discrimination), as they may play a role in the development of MLTCs in different ethnic groups.

## BACKGROUND

Long□term conditions (e.g. chronic kidney disease, hypertension and depression) are health conditions that are currently not curable and can only be managed with medication or other therapies [1, 2]. One in four primary care patients in the United Kingdom (UK) have multiple long-term health conditions (MLTCs) i.e. the presence of two or more long-term health conditions in an individual [3, 4]. Further, the proportion of people living with four or more long-term conditions is expected to double between 2015 and 2035 [5]. Evidently, healthcare systems, which have previously focused on single conditions, will need to radically transform their approaches to meet the challenges and complexity of caring for people with MLTCs [6].

Indicative evidence suggests that the risk of developing MLTCs is higher, and with MLTCs occurring at an earlier age, for people from minoritised ethnic groups than people from the majority White group [7, 8]. Studies of single conditions provide evidence that many, but not all, people from minoritised ethnic groups in the UK experience poorer health than people from the White ethnic group [9, 10]. Much of the variation in poor health across ethnic groups is due to underlying socioeconomic inequalities which, in turn, can be attributed to life-course experiences of racism and racial discrimination [11].

With ethnic inequalities evident in single health conditions, we would expect ethnic inequalities in MLTCs to follow a similar pattern. To our knowledge, there has been one previous review that has reported on ethnicity and MLTCs in the UK [12]. However, this review narrowly focused on long-term mental health conditions (i.e. comorbid substance use in psychosis), and therefore, provides only partial evidence of the burden of MLTCs across ethnic groups. Given that having MLTCs is associated with poorer functioning, lower quality of life, higher mortality risk [4], and greater healthcare use and cost [13], developing an understanding of the variation in the burden of MLTCs across ethnic groups in the population is key to ensure efficient, and equitable policy and practice [7]. Thus, the aim of the present review is 1) to identify and describe the literature that provides evidence of ethnicity and prevalence of MLTCs amongst people living in the UK, and 2) to summarise the prevalence estimates of MLTCs across ethnic groups.

## METHODS

### Search strategy

As per the Preferred Reporting Items for Systematic review and Meta□Analysis Protocols (PRISMA□P) guidelines [14], we registered the protocol for this review on PROSPERO (CRD42020218061). We electronically searched for studies that included the prevalence of MLTCs across ethnic groups in the UK using the following databases: ASSIA (Applied Social Sciences Index and Abstracts), Cochrane Library, EMBASE (Excerpta Medica dataBASE), MEDLINE, PsycINFO, PubMed, ScienceDirect, Scopus, Web of Science core collection. To ensure that relevant grey literature was not excluded we also conducted a search on OpenGrey. We supplemented the electronic search with a manual search of the reference lists of colleagues working in the field and of key articles identified. When full texts were unavailable, we contacted the relevant authors.

We adhered to the conventions of each search engine and used search terms that captured the key concepts; Ethnicity (e.g. “Ethnic Groups”[Mesh] OR “BME” OR “BAME”), multiple health conditions (e.g. “Multiple Chronic Conditions” OR Comorbid* OR Multimorbidity), Health inequality (e.g. “Health Equity”[Mesh] OR “Healthcare disparit*” [MeSH] OR Inequalit*) and the geographical location (e.g. “United Kingdom”[MeSH Terms] OR “UK”) (See Supplementary file 1 for a full list of search terms).

### Study inclusion and exclusion criteria

We did not restrict the start of the search to any particular period in time and included studies published up until December 2020. Only UK studies, reported in English, with estimated prevalence of MLTCs across ethnic groups of people in the UK general population, residing in the community were included. To accommodate the variety of ways in which MLTCs were defined and operationalised in the extant literature, we included studies of multimorbidity (i.e. the presence of two or more long-term health conditions[15]) and comorbidity (i.e. the presence of any distinct additional co-existing ailment in a person with an index condition under investigation[16]). We applied further restrictions to address the second objective (to summarise the prevalence estimates of MLTCs across ethnic groups). Given the role of age in patterning MLTCs [2], and the younger age profile of minoritised people[17], we excluded studies that did not adjust for at least age as they would have provided an inaccurate representation of the prevalence of MLTCs across ethnic groups. We also excluded studies that focused on only two conditions (e.g. depression and substance abuse) and included only studies that counted more than two conditions. These studies are more likely to focus on people with overall severity of illness, greater healthcare utilisation, and complex medical needs [18, 19].

We imported the studies retrieved from the electronic search to Endnote X8 where duplicates were removed. Two reviewers, BH and LB, screened a random sample (10%) of the titles, abstracts, and full texts. BH independently screened the remaining studies. Disagreements were resolved by discussion.

### Data extraction strategy

We extracted relevant data from the included studies using a structured form with the following items: study identifier, design, setting, recruitment/data source, sample size, population description, definitions of MLTCs, type and number of MLTCs, confounders, and the results. BH and LB extracted data from a random sample (10%) of the included studies. Differences were reconciled through discussion. BH independently extracted data from the remaining studies.

### Population and Outcomes

The outcomes of interest were prevalence estimates for MLTCs for at least one minoritised ethnic group, compared to the White majority population.

### Data synthesis and presentation

Given the different ways in which MLTCs were conceptualised and operationalised, the different ethnic groups, and the range of conditions explored in the included studies, we conducted a narrative synthesis. We present the findings of the synthesis in themes, supplemented with tables and figures, in two sections. First, we provide an overview of the studies that documented the prevalence estimates of MLTCs across ethnic groups. Second, we present studies where the authors counted more than two long-term conditions and reported on at least age-adjusted prevalence estimates of MLTCs across ethnic groups. We use the terminology used by authors to describe ethnic categories in their studies.

## RESULTS

### Overview of included studies

We identified 7677 titles from the electronic search (See Figure 1 which is based on PRISMA guidelines [20]). After removal of duplicates and studies identified as ineligible from the title or abstract, 188 papers were eligible for further evaluation. A further 104 studies were excluded, producing a final sample of 84 studies for the review. Seven of these studies contributed to the evidence of ethnic inequalities in the prevalence of MLTCs among people living in the UK. These were studies in which the authors counted more than two long-term conditions and adjusted for at least age in their analyses.

**Figure 1:**
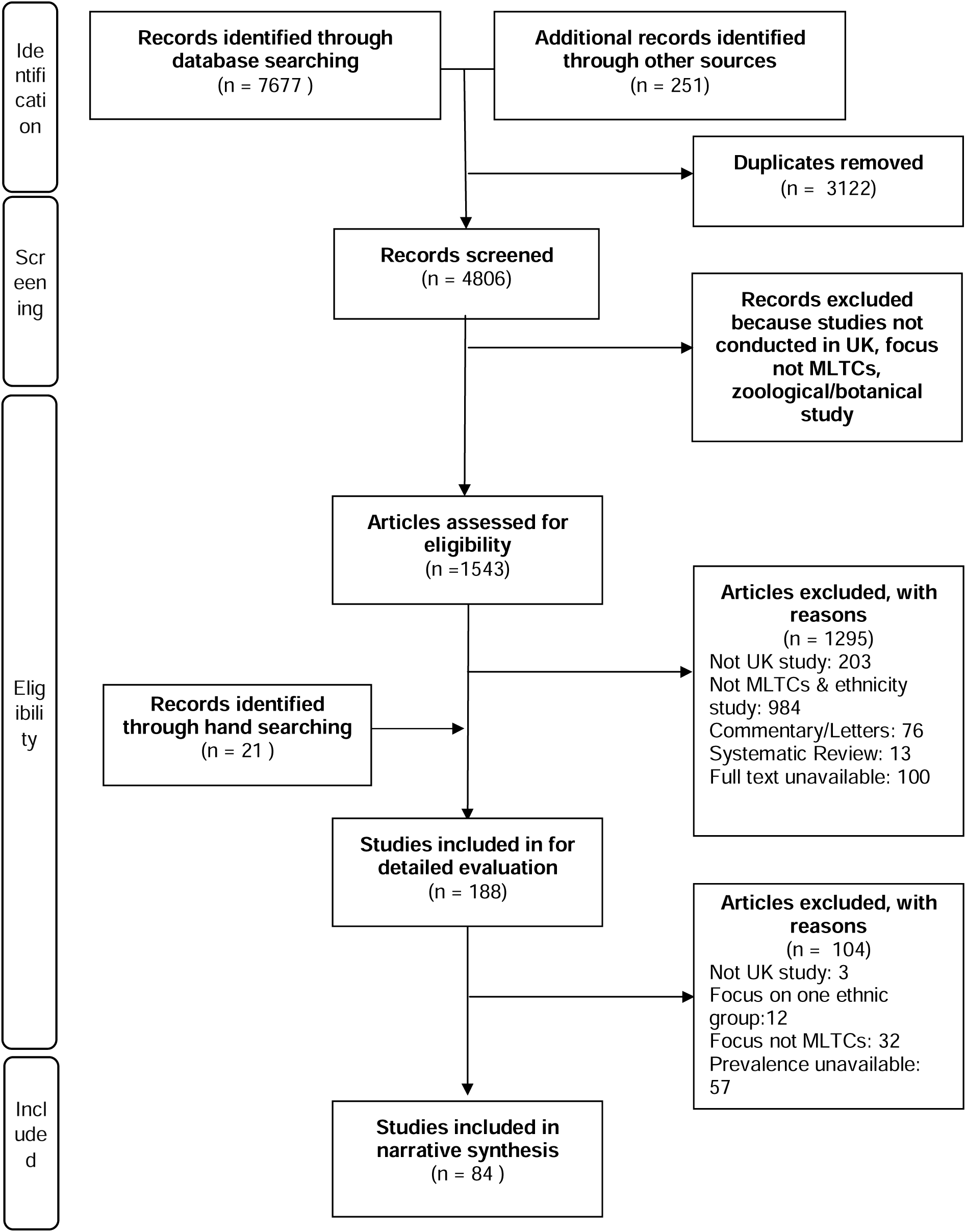
PRISMA Flow chart.

The 84 studies included were published between 1984 and 2020. 49 studies were conducted locally, 7 studies were regional, and 28 were national. Sample sizes ranged from 45 to nearly 900,000. The majority of the studies used patient records to analyse the prevalence of MLTCs in people from minoritised ethnic groups (n=68) (Figure 2). These studies used data from primary care (n=19), hospital records (n=14), specialist clinics/services (n=19) and disease registers (n=17). Fourteen used cross-sectional survey data (e.g. the General Practice Patient survey, Mental Health and Substance Misuse services survey, and the National tuberculosis surveys) and cohort study data (e.g. the HUSERMET Study, the Yorkshire Health Study, the Southall and Brent Revisited study, the Comorbidity Dual Diagnosis Study (n=2), the Millennium Cohort Study (n=2), and UK Biobank (n=4)) [21-34]. In one study, the authors used Scotland-wide linked education and patient databases [35].

**Figure 2:**
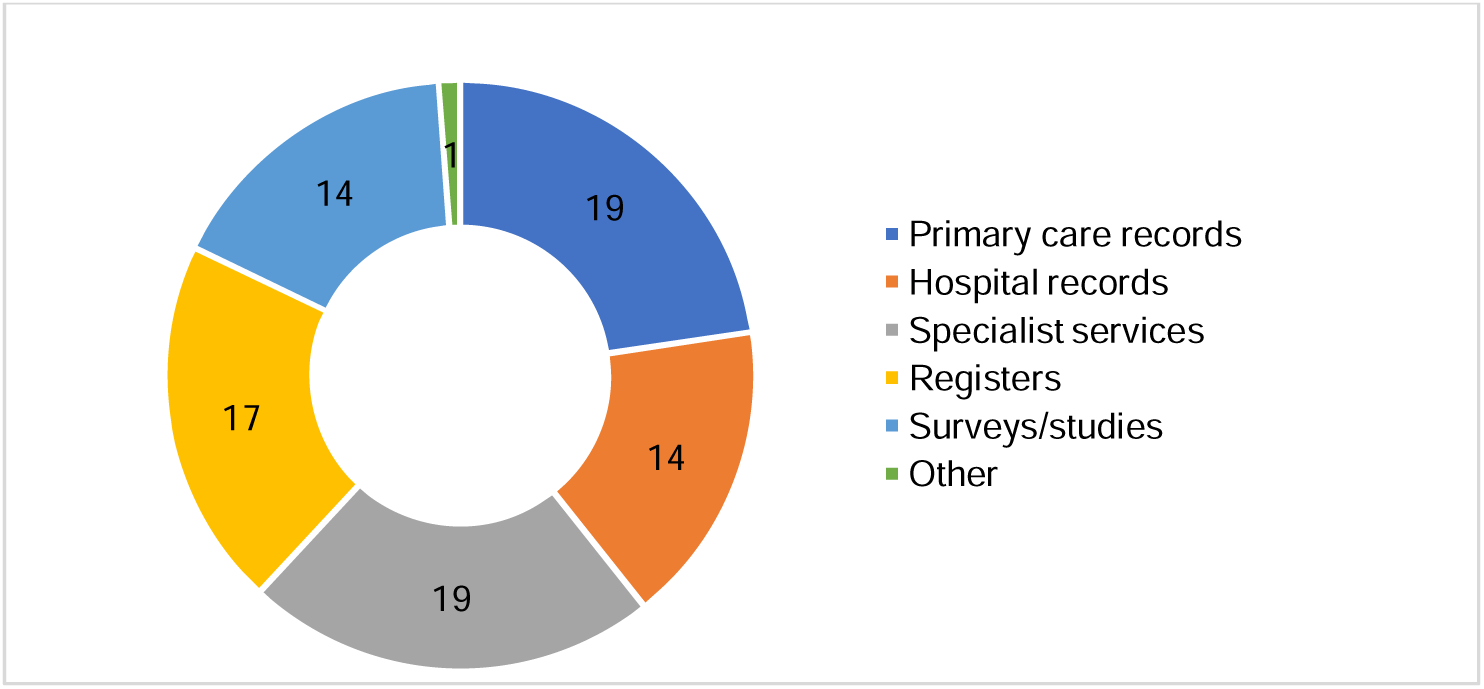
Data Sources used in included studies.

### Population characteristics

#### Ethnic group identification

38 studies (45%) explicitly reported how ethnicity was identified. Of these, participants self-reported their ethnic identity in 23 studies (27%). One of these studies determined ethnicity based on genealogy [26]. In four studies ethnicity was assigned by interviewers [36], keyworkers [34], researchers, physicians, and nurses [37-39]. In six studies, ethnicity was both self-reported by participants and clinician assigned [40-45]. Four studies used computerised name recognition programs to identify South Asian people [46-49]. In one study, ethnicity was identified using self-reports, specialist unit ascription, and name recognition software [49].

When ethnicity data could not be identified, some authors attempted to locate the missing information in a variety of ways. One study obtained missing ethnicity data from hospital services and primary care records [50]. Another study imputed the missing participants’ ethnicity from the Census super output area using postcode of residence; where this was an area with ≥ 98% White ethnicity, they assumed the participant was of white ethnicity [49]. The authors in one study relied on the modal ethnicity of patients with the same surname in the Electronic Health Records database where possible [51].

#### Ethnic group categorisation

Of the 84 included studies, 60 compared the prevalence of MLTCs across four ethnic categories or fewer. In 13 studies participants were grouped into five categories. A further 11 studies grouped their participants into six categories or more (Supplementary file 2). Table 1 below provides a breakdown of the ethnic groups of interest in the 18 studies that compared the prevalence of MLTCs between two ethnic groups. Two studies which compared particular minoritised ethnic groups (i.e. Indian and Punjabi people) with White European and English people [52, 53]. The rest used broad minoritised group categories. The majority (n=12) compared Asians/South Asians with White/British/Caucasian/European participants [33, 46-48, 52-59]. Two studies categorised participants into Black/Afro-Caribbean and White/Caucasian [60, 61]. Four studies did not specify a minoritised group and categorised participants into either White/Caucasian or non-White/other [23, 29, 32, 62].

**Table 1.** Ethnic groups in studies comparing 2 ethnic group categories.

| **Asian/South Asian ethnicity compared with majority white ethnic group** |  |
| --- | --- |
| Asian | British[54] |
| Asian | White[55, 56] |
| Asian | Caucasian[57] |
| Indian | European[52] |
| Punjabi | English[53] |
| South Asian | Caucasian[58] |
| South Asian | European[33] |
| South Asian | Europid[59] |
| South Asian | White European[47, 48] |
| South Asian | Non-South Asians[46] |
| **Black/African/Caribbean ethnicity compared with majority white group** |  |
| Afro-Caribbean | Caucasian[61] |
| Black | White[60] |
| **Unspecified ethnicity compared with majority white ethnic group** |  |
| Non-Caucasian | Caucasian[32] |
| Non-white | White[23, 29] |
| Other | White[62] |

In some studies, ethnic categories with small numbers were excluded from the analyses [60, 63] [21, 49] [64, 65].

#### Missing ethnicity data

The percentage of missing ethnicity data was available in 25 studies (30%). Some studies excluded participants with missing ethnicity data from the analyses [66, 67] or reported these as ‘unknown’[37]. Only four studies conducted sensitivity analyses to ascertain if there were differences between missing cases and complete cases [25, 31, 64, 68].

#### Age

The participants’ age was reported in 81 studies. There were 75 studies that included participants aged 16 and above. Of these studies, 31 reported a mean age of 40 years and above. Three studies reported a mean age of between 30 and 39 years [28, 36, 69]. Four studies investigated the prevalence of MLTCs in children [31, 32, 35, 70], and two studies included both adults and children [71, 72].

### Studies reporting on ethnicity and prevalence of MLTCs (Unspecified Index condition)

Of the 84 studies, nine studies counted the number of long-term conditions where there was no specific index condition. The number of ethnic group categories varied across the studies (Table 2).

**Table 2.** Studies reporting MLTCs across ethnic groups with no index condition.

| Study ID | Study design | Geographical reach | Data Source | Sample size | Participant characteristics (Age in years) | Ethnic categories | Index condition | Covariates adjusted for in analysis of the prevalence of multiple conditions |
| --- | --- | --- | --- | --- | --- | --- | --- | --- |
| Ashworth et al., 2019 [73] | Longitudinal study | Local | Primary care records | 332353 | %Female:50;<br>Age:≥18 | White, Black, South Asian, Mixed, Other | n/a | age, gender, deprivation, cardiovascular risk |
| Chudasama et al., 2020 [23] | Longitudinal cohort study | National | UK Biobank | 480,940 | %Female: 54;<br>Age range: 38-73 | White, Non-White | n/a | none |
| Dorrington et al., 2020 [74] | Longitudinal study | Local | Lambeth Datanet | 326415 | %Female:53;<br>Age range: 16-60 | White, Black African, Asian, Black Caribbean, Other mixed, Other, Black other | n/a | age, gender, deprivation |
| Fleming et al., 2020 [35] | Retrospective study | National | Scotland-wide databases | 766,244 | %Female: 49%;<br>Age range: 4-19 | White, Asian, Black, Mixed, Other | n/a | none |
| Hesketh et al., 2016 [31] | Longitudinal study | National | Millennium Cohort Study | 9548 | %Female:51;<br>Age <18 | White, Mixed, Indian, Pakistani/Bangladeshi, Black, Other | n/a | Sex, highest level of maternal education, quintiles of household income |
| Li et al.,2016 [29] | Longitudinal study | Local | Yorkshire Health Study. | 27806 | %Female:56;<br>Age range:16-85 | White, non-White | n/a | none |
| Mathur et al., 2011 [66] | Cross-sectional study | Local | Primary care records | 99648 | %Female: NR;<br>Age: ≥18 | White, South Asian, Black, Other | n/a | age, sex |
| Paddison et al., 2015 [25] | Cross-sectional study | National | General Practice Patient Survey | 890427 | %Female:56;<br>Age: ≥18 | White, Mixed, Asian, Black, Other | n/a | none |
| Zemedikun et al., 2018 [22] | Cross-sectional study | National | UK Biobank | 502643 | %Female:54;<br>Median Age(IQR): 58(50-63) | White, Mixed, Asian, Black, Other | n/a | none |

### Studies reporting on ethnicity and prevalence of MLTCs (Index condition specified)

The majority of studies reporting MLTCs across ethnic groups were studies that specified an index condition (n=75) (Table 3). Measures of comorbidity were used in five studies; three of which used the Charlson Comorbidity Index [72, 81, 99], one used the Elixhauser Comorbidity score [37], and another used the Stoke comorbidity score [84]. As shown in Table 3, diabetes (n=14), renal disease (n=14), and psychotic illness (n=7) were the most frequently studied index conditions. COVID-19 was the index condition in three studies [37, 51, 67].

**Table 3.**
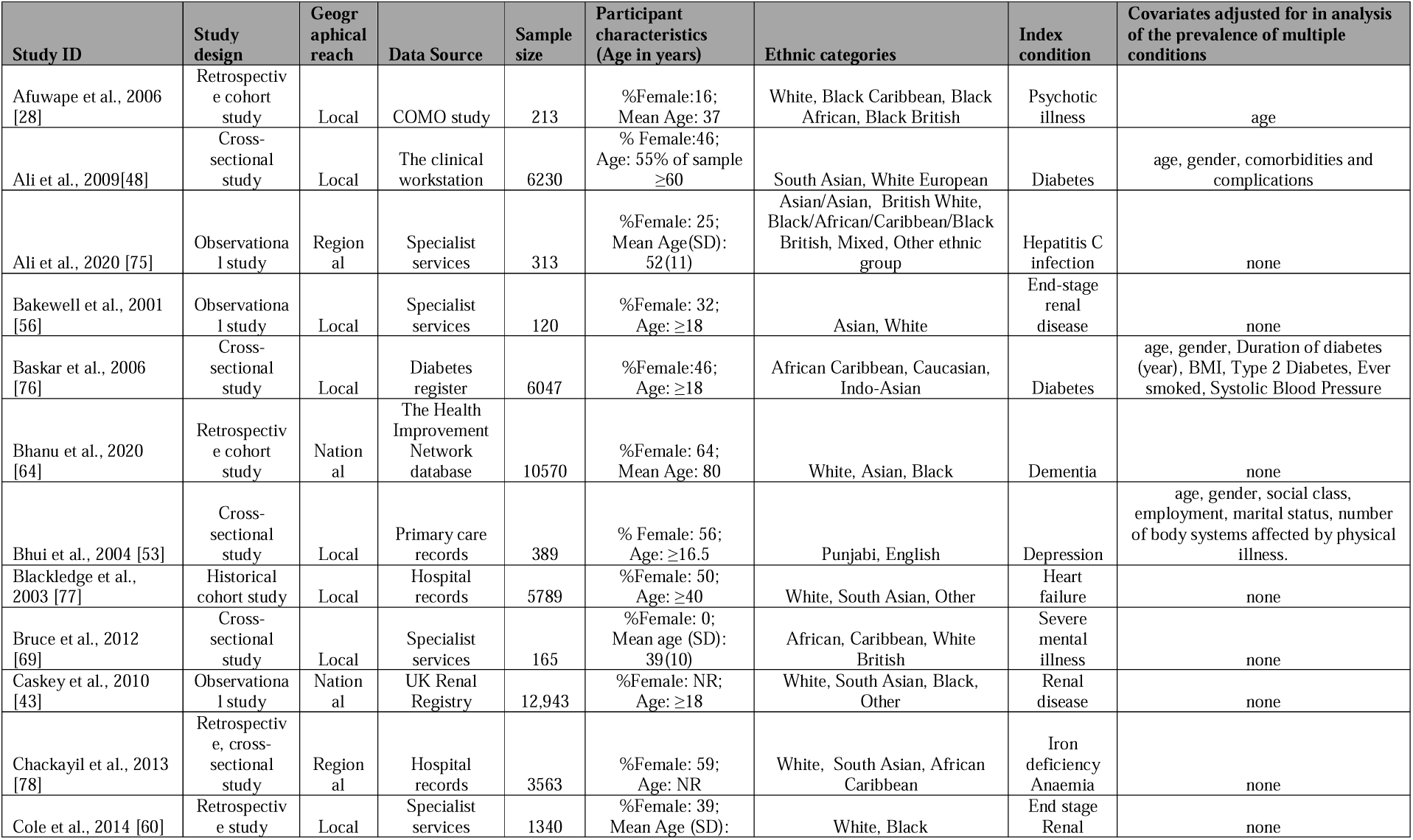
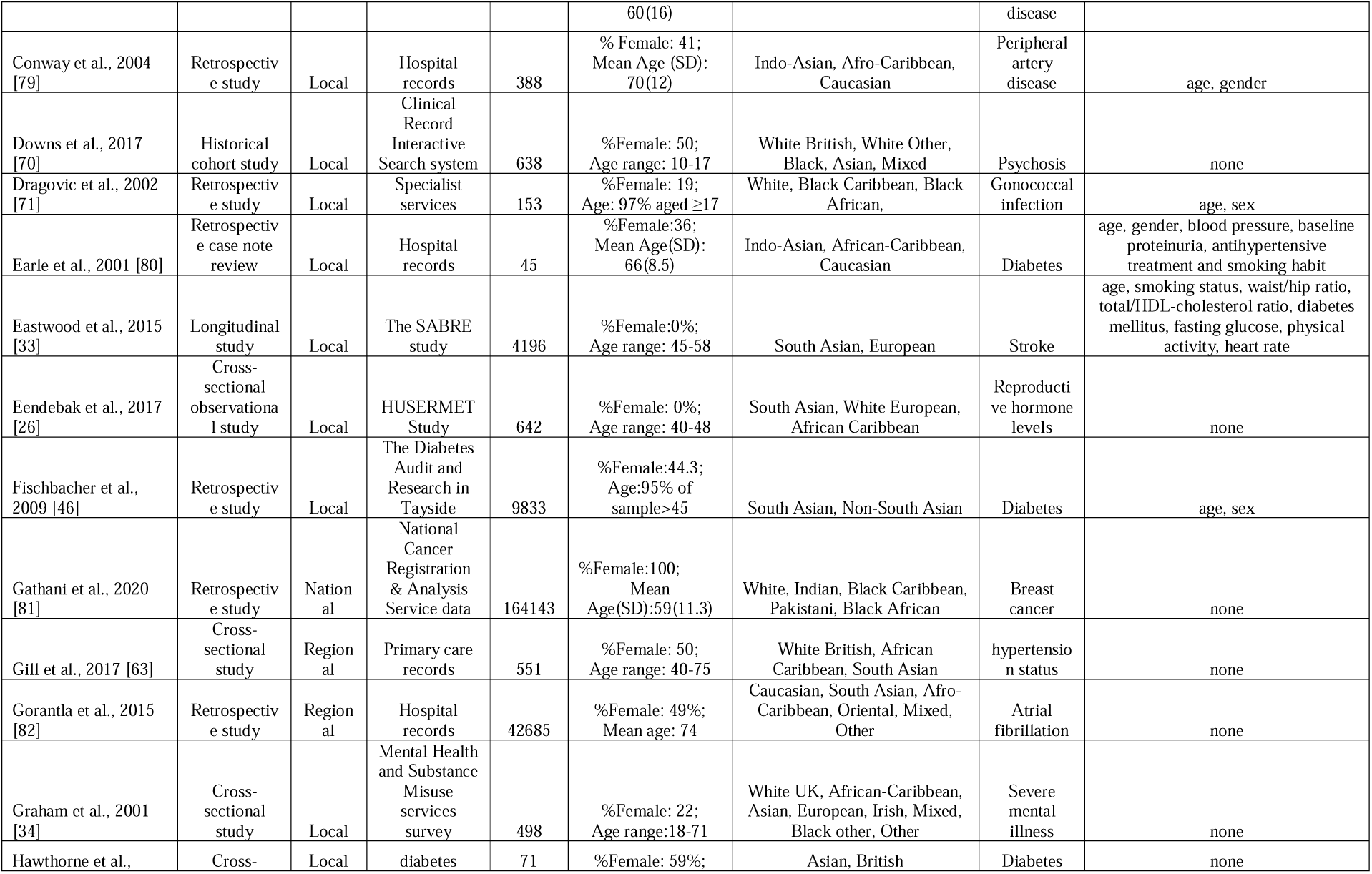
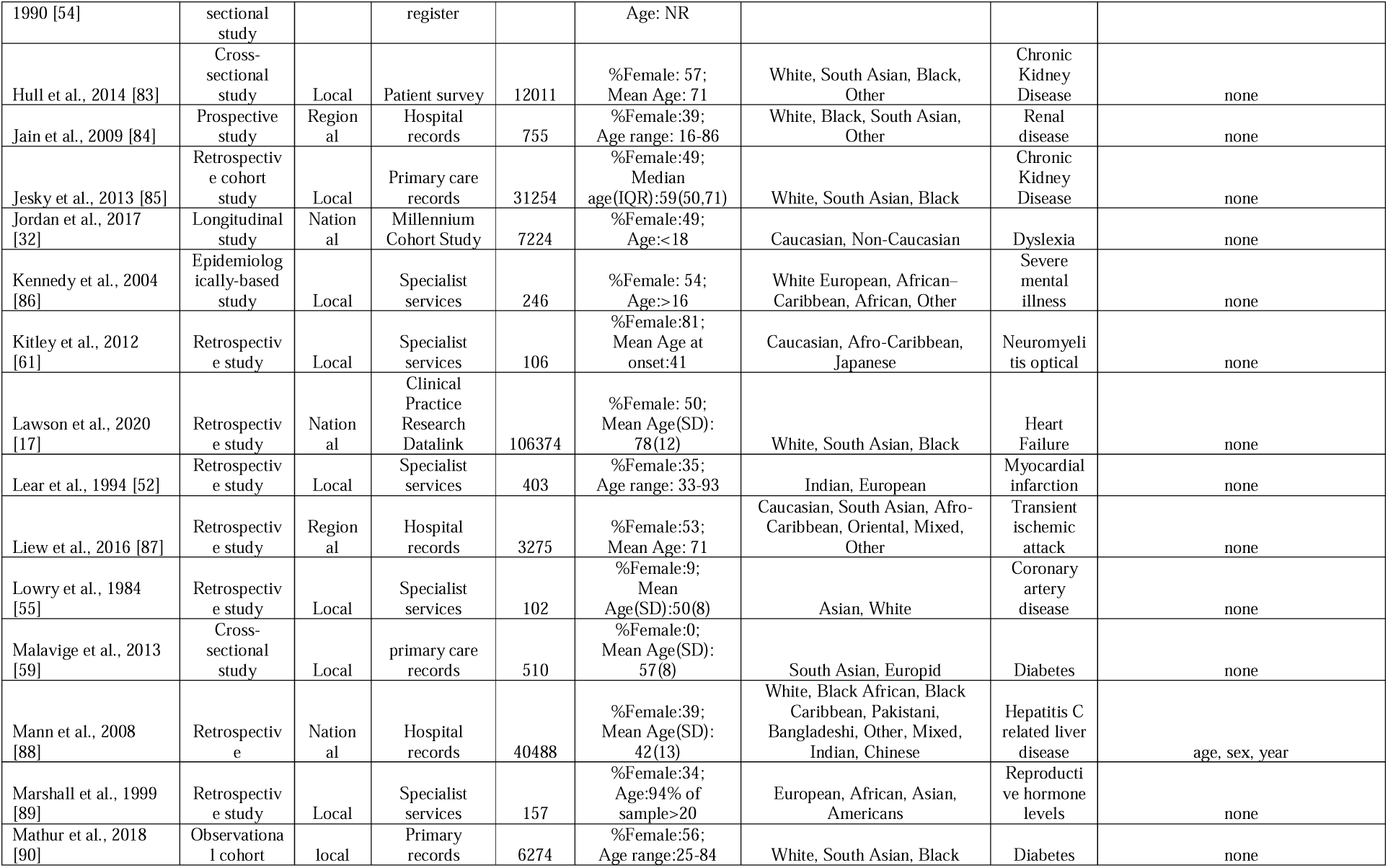
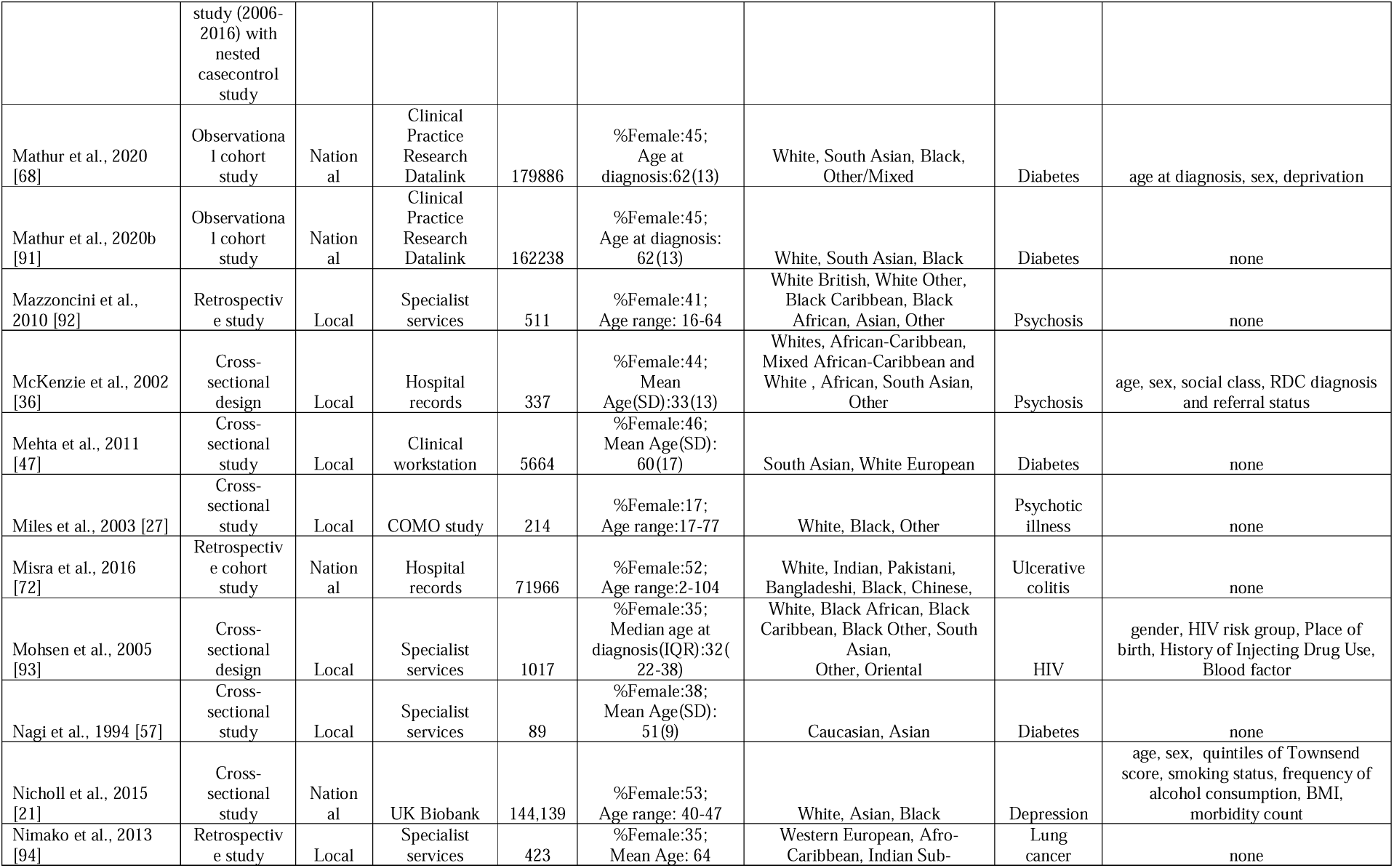
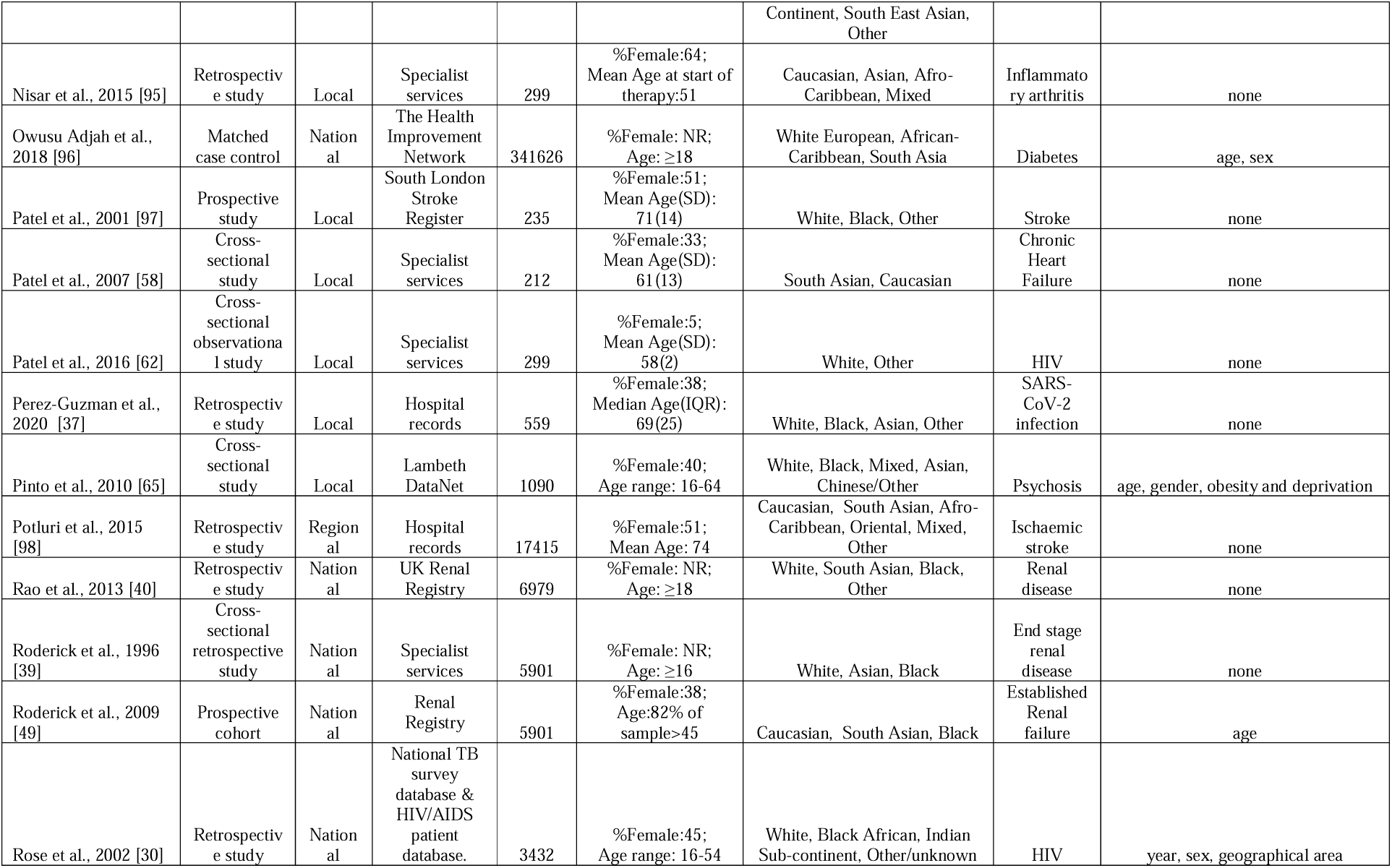
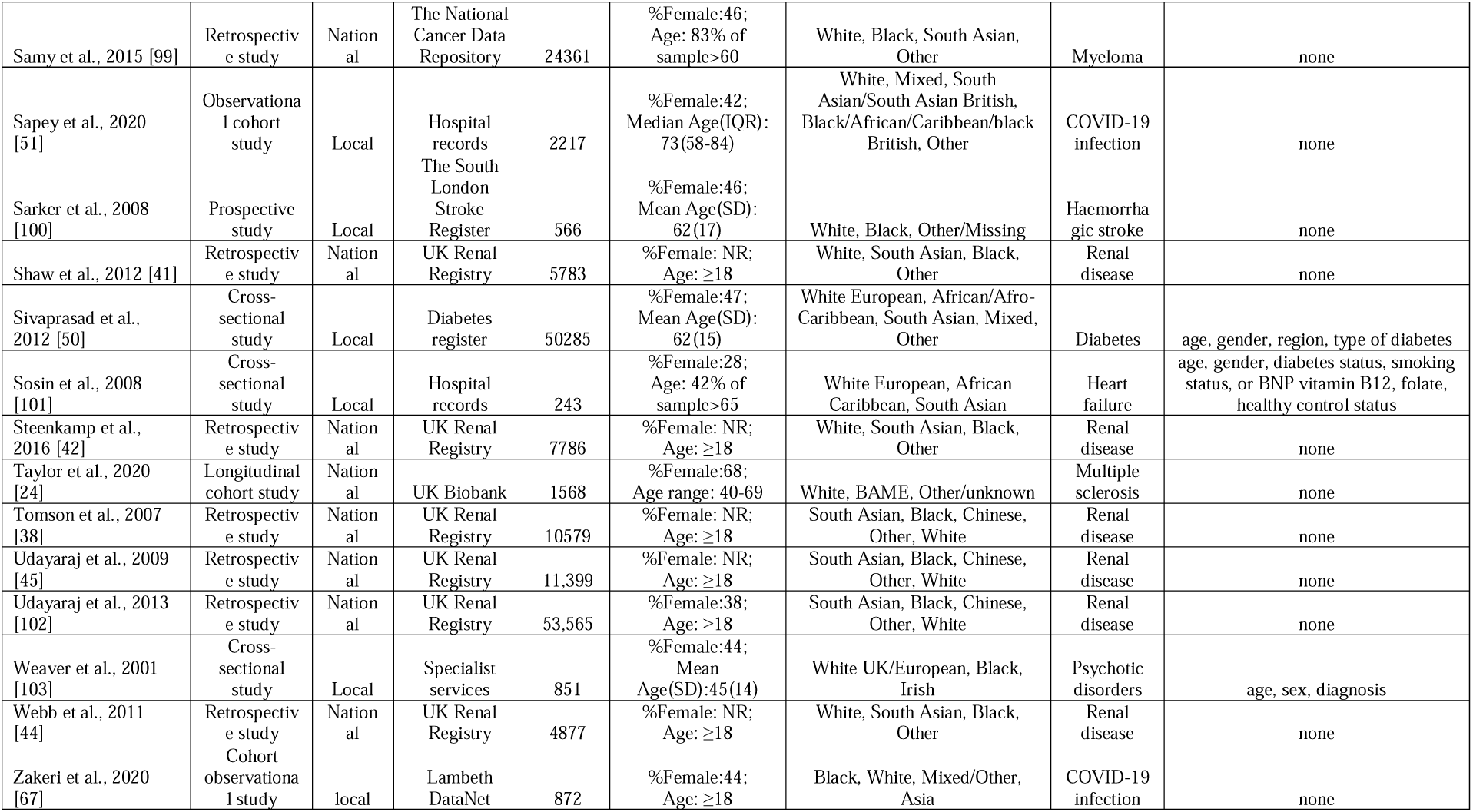
Studies reporting MLTCs across ethnic groups specifying an index condition.

### Prevalence of multiple long-term conditions by ethnic group

Seven studies provided evidence on ethnicity and age-adjusted prevalence of multiple conditions amongst people living in the UK (Table 4). The studies are highlighted to denote whether the results suggest minoritised ethnic groups have higher (↑), or lower (↓) levels of MLTCs when compared with the White majority group. Of these, three were studies of MLTCs with no specified index condition [66, 73, 74]. The findings of these studies suggest that people from minoritised ethnic groups have a higher prevalence of MLTCs when compared to people from the White majority group [66, 73, 74]. All were local studies that used patient records in their analyses. All adjusted for age and gender/sex, with two studies also adjusting for deprivation [73, 74] and one also adjusting for cardiovascular risk factors [73]. Mathur and colleagues investigated the cardiovascular multimorbidity by ethnicity in a socially deprived and multi-ethnic population in east London [66]. Their focus was on five health conditions: coronary heart disease, diabetes, heart failure, stroke, and hypertension across four ethnic groups. After adjusting for age and sex, and clustering by general practice, minoritised ethnic groups were more likely to be multimorbid than those who were White with adjusted odds ratios of 2.4 (95% Confidence interval (CI): 1.94-2.15) for South Asian people and 1.23 (95% CI: 1.18-1.29) for Black people [66].

**Table 4.** Studies reporting adjusted prevalence of MLTCs multiple conditions across ethnic groups.

| Study ID | Region | Data Source | Sample size | No. of Ethnic Categories | No. of conditions | Types of conditions | Index condition | Covariates | Higher/Lower Prevalence of MLTCs |
| --- | --- | --- | --- | --- | --- | --- | --- | --- | --- |
| Mathur et al., 2011 | Local | Primary care records | 99648 | Black, Other, South Asian, White | 5 | Hypertension, Ischaemic Heart Disease, Heart Failure, Stroke, And Diabetes | - | age, sex | ↑ |
| Ashworth et al., 2019 | Local | Primary care records | 332353 | Black, Mixed, Other, South Asian, White | 12 | Atrial Fibrillation, Chronic Obstructive Pulmonary Disease , Chronic Pain , Chronic Kidney Disease, Coronary Heart Disease , Diabetes Mellitus , Dementia, Depression, Heart Failure, Serious Mental Illness, Stroke, Morbid Obesity | - | age, gender, deprivation, cardiovascular risk | ↑ |
| Dorrington et al., 2020 | Local | Lambeth Datanet | 326415 | Asian, Black African, Black Caribbean, Black Other, Mixed, Other, White | 13 | Epilepsy, Chronic Pain, Depression, Vascular, Cancer, Cardiac Disease, Rheumatoid Arthritis, Severe Mental Illness, Hypertension, Diabetes, Obesity, Respiratory Disease, Learning Disability | - | age, gender, deprivation | ↑ |
| Roderick et al., 2009 | National | Renal Registry data | 5901 | Black , South Asian, White | 8 | Established Renal Failure, Coronary Heart Disease, Diabetes, Any Vascular Disease, Chronic Obstructive Pulmonary Disease, Smoking, Any Malignancy, Chronic Liver Disease | Established Renal Failure | age | ↑ |
| Baskar et al., 2006 | Local | Diabetes Register | 6047 | Afro-Caribbean, Caucasian, Indo-Asian | 6 | Diabetes, Retinopathy, Dipstick Proteinuria, Ischaemic Heart Disease, Cerebrovascular Disease, Peripheral Vascular Disease | Diabetes | age, gender, diabetes complication risk factors | ↑ |
| Owusu Adjah et al., 2018 | National | THIN data | 341626 | African-Caribbean, South Asian, White European | 5 | Diabetes, Myocardial Infarction, Heart Failure, Stroke, Cancer | Diabetes | age, sex | ↓ |
| Mathur et al., 2020 | National | CPRD data | 179886 | Black, South Asian, White | 8 | Diabetes, Hypertension, Coronary Heart Disease (Angina/Myocardial Infarction), Heart Failure, Chronic Kidney Disease, Retinopathy, Neuropathy | Diabetes | age, sex, deprivation | ↓ |

↑prevalence of MLTCs is higher in minoritised people; ↓prevalence of MLTCs is lower in minoritised people; THIN-The Health Improvement Network; CPRD- Clinical Practice Research Datalink;

Ashworth and colleagues examined the social determinants and cardiovascular risk factors for multimorbidity and the acquisition sequence of multimorbidity [73]. Their focus was on 12 conditions across five ethnic groups. Age-sex adjusted estimates were not provided but they found that Black and South Asian people had higher odds of multimorbidity after adjusting for age, gender, and area level deprivation (OR 1.15 (95% CI: 1.07-1.23) and 1.19 (95% CI: 1.07-1.33) respectively). However, when hypertension, obesity, and smoking status were added to the model, only South Asian people had higher odds of multimorbidity (1.44 [95% CI: 1.29-1.61]), but not Black people (0.86 [95% CI: 0.80-0.92]) [73].

Dorrington and colleagues assessed 13 conditions across seven ethnic groups. Age-sex adjusted estimates were not provided [74]. Age, gender and area-level deprivation adjusted models showed that the odds of having two or more long-term conditions were higher in some minoritised ethnic groups (1.92 (95% CI: 1.75–2.10) for Black African people, 2.83 (95% CI: 2.56–3.14) for Black Caribbean people, 1.50 (95% CI: 1.31–1.72) for people with Mixed ethnicity and 2.22 (95% CI: 1.87–2.64) for people who self identified as Black Other. When three or more long-term conditions were considered, the same minoritised ethnic group populations still had higher odds of multimorbidity than the White population (OR 3.42 [95% CI:3.05–3.83] for Black Caribbean people, 2.53 [95%CI: 2.02–3.18] for Black Other people, 1.95 [95% CI 1.74–2.20] for Black African people and 1.50 [95% CI: 1.30–1.73] for people with Mixed ethnicity) [74].

In the remaining four studies, an index condition was specified [49, 68, 76, 96]. Two studies, one which focused on people starting renal replacement therapy, and the other which focused on people with diabetes, reported a higher prevalence of MLTCs in people from some minoritised ethnic groups [[76]48]. The other two studies which focused on people with diabetes, reported lower prevalence of MLTCs among minoritised people [68, 96]. Three out of these four studies used national data [49, 68, 96]. All used patient records to assess MLTCs across three ethnic group categories: Black/African-Caribbean, South Asian/Indo-Asian and White ethnicity. All four studies adjusted for age, three studies additionally adjusted for sex [68, 76, 96]. One study additionally adjusted for area-level deprivation [68] and another for diabetes complication risk factors [76].

In their study exploring the survival of patients starting renal replacement therapy across three ethnic groups, Roderick and colleagues found that compared to White patients, the age-adjusted prevalence of vascular comorbidity was higher in South Asian patients but lower in Black patients [odds ratios of 1.26 (95% CI:1.04– 1.52) and 0.70 (95% CI:0.52–0.95) respectively]. However, other comorbidities were found to be generally common in White patients [49]. Baskar and colleagues evaluated ethnic differences in the prevalence of hypertension and vascular complications in a population with diabetes[76]. They considered microvascular complications to be the documented presence of any grade of retinopathy and/or dipstick proteinuria [76]. Macrovascular complications were considered to be the documented presence of ischaemic heart disease and/or cerebrovascular disease, and/or peripheral vascular disease [76]. Age-adjusted estimates were not provided. After adjusting for age, gender, BMI, systolic blood pressure, smoking, type 2 diabetes, and duration of diabetes, Afro-Caribbean people had a higher risk of microvascular complications (odds ratios of 1.293 (95% CI: 1.063–1.573) relative to White people). However, compared to White people, both Afro-Caribbean and Indo-Asian people had significantly lower risk of macrovascular complications with odds ratios of 0.710 (95% CI: 0.581–0.866) and 0.807 (95% CI: 0.669–0.933) respectively [76].

Owusu-Adjah and colleagues examined ethnic differences in comorbidities in patients with type 2 diabetes mellitus [96]. They found that Western European patients had significantly higher baseline age-sex adjusted prevalence of cardiovascular complications compared to South Asian patients (at all levels of BMI) and African-Caribbean patients (in overweight or obese groups only). Western Europeans also had significantly higher baseline prevalence of cancer and depression [96]. Mathur and colleagues investigated ethnic differences in the severity and clinical management of type 2 diabetes at initial diagnosis. Age-adjusted estimates were not provided. Adjusting for age, sex and deprivation, and clustering by practice, the odds of having comorbid macrovascular disease (i.e. hypertension, coronary heart disease (including myocardial infarction and angina), stroke, and heart failure) at diagnosis were reduced in South Asian people (0.88, 95%CI 0.80–0.96) and halved in Black people (0.50, 95%CI 0.43–0.58) relative to White people. However, they found no ethnic differences in the odds of having diagnosed microvascular disease (i.e. chronic kidney disease, retinopathy, and neuropathy) in their sample [68].

## DISCUSSION

We identified seven studies that give insight into age-adjusted ethnic differences in the prevalence of MLTCs. The findings are indicative of ethnic inequalities in MLTCs in favour of the White British majority as five of the seven studies reported that some minoritised ethnic groups have a higher prevalence of MLTCs than their White counterparts. The evidence suggests that South Asian people (three out of five studies) and Black people (two out of five studies) may be at a higher risk of MLTCs [49, 66, 73, 74, 76]. Whilst some studies adjusted for factors that may be on the explanatory pathway, including deprivation and risk factors for cardiovascular disease and diabetes complications, all studies adjusted for at least age in their analyses. As such, the evidence of ethnic inequalities in MLTCs is based on studies that considered this key confounder [2]. However, given the variation in the number and types of conditions examined in these studies and the merging of different minoritised ethnic groups, this evidence may not accurately reflect the true level of inequality.

The two studies that reported a lower prevalence of MLTCs in minoritised ethnic groups both focused on people living with diabetes. This finding is intriguing and warrants further attention. In trying to understand these observations, we must consider that Black and South Asian people in the UK not only have a higher prevalence of diabetes, but they also develop the condition at an earlier age [91, 104]. It is, therefore, possible that the minoritised populations included in these studies are younger than their White counterparts. Since MLTCs increase with age, the lower prevalence in MLTCs among minoritised ethnic groups observed in these studies could result from residual confounding by age. Also, given that minoritised ethnic groups have a higher risk of developing diabetes, much effort might be paid to the identification and management of diabetes in this population. Mathur and colleagues, who examined the ethnic variations in the severity and management of diabetes at first diagnosis, provide support for this notion [68]. They found that when compared to White people, Black and South Asian people had better capture of risk factors, better/similar cardio-metabolic profile at diagnosis, faster initiation of anti-diabetic treatment, first National Health Service (NHS) health check and structured education [68]. These outcomes may prevent further health problems, thereby contributing to a lower prevalence of MLTCs in minoritised people with diabetes. However, other studies report ethnic inequalities in diabetes care with Black and Asian people found to have worse glycaemic control and being less likely to be prescribed newer therapies [105]. Inequalities in diabetes care can result in higher rates of complications [105] which can increase the likelihood of MLTCs. Future work is, therefore, required to explore these findings further.

### Sources of data

All seven studies analysed data from patient records. For individuals to be included in patient records, they need to be in contact with healthcare services. It has been reported that access to primary care health services is generally equitable for people from minoritised ethnic groups [106]. However, they are less likely to access other specialist services [106, 107]. Therefore, those who do not access these services or for that matter, those who do not use healthcare services at all, are excluded from analyses that employ patient records. This differential access may lead to underestimation of inequalities. It is also important to consider ethnic inequalities in care quality as studies suggest that there may be differences in how patient symptoms are recorded, diagnosed or treated [108]. Other studies have not only found data quality problems when ethnicity data is recorded in hospital records (e.g. incomplete coding, inconsistent use of codes, systematic biases), but also that these data quality issues disproportionately affect hospital records for minoritised patients [109]. Consequently, the health conditions in some minoritised people may be underreported when patient records are used in analyses, thereby, impacting on the prevalence estimates of MLTCs and impeding our understanding of ethnic inequalities.

### Broad ethnic group categories

In the seven studies that contributed to the evidence of ethnic inequalities in the prevalence of MLTCs, people from minoritised ethnic groups were often grouped into broad categories; in particular, Black/African-Caribbean, South Asian/Asian/Indo-Asian, Mixed, and Other. In one study, the Black ethnic group was disaggregated into three different categories, but the authors did not adopt this approach for the Asian ethnic group [74]. Categorising minoritised ethnic group people into overarching categories can be useful for identifying broad patterns as some may have shared experiences of racism, discrimination, and/or social exclusion [28]. However, the use of broad ethnic categories may obscure the extent of inequalities. For example, as reported above, South Asian people may be at particular risk of MLTCs [49, 66, 73] but studies have shown that Pakistani and Bangladeshi women report higher levels of limiting long-term illness than Indian women [9].

Yet, they are often grouped together as people of South Asian ethnicity. This issue also applies to the White other ethnic group [36] who are a diverse population with particular groups, e.g. Gypsy, Roma, and Traveller community [9, 110].

### Missing ethnicity data

The availability of complete ethnicity data is crucial for studies that seek to assess ethnic inequalities not only in relation to MLTCs, but also in relation to healthcare utilisation and care[111]. Information about the different ways in which authors handled missing ethnicity data was available in all but one of the seven studies that contributed to the evidence of ethnic inequalities in the prevalence of multiple conditions [74]. Amongst these studies, people with missing ethnicity data were excluded from analyses in three studies and combined with the ‘other’ ethnic group in one study [66, 73, 96]. Of note is that the authors of only one of these six studies reported that they conducted sensitivity analysis; comparing individuals of known ethnicity to those whose ethnicity data was missing [68]. Researchers should carefully consider the most appropriate approach to dealing with missing data and check alternative approaches using sensitivity analysis because policies based on inaccurate data may result in poor targeting of resources and services [111].

### Proposed mechanisms underlying observed inequalities

Given that few studies have assessed the prevalence of MLTCs across ethnic groups, it is difficult to ascertain the reasons behind the observed ethnic inequalities in the prevalence of MLTCs in the UK. However, based on longstanding international evidence on ethnic inequalities in single conditions, we propose a number of mechanisms. We consider the impact of racism and discrimination in explaining the observed ethnic differences in the prevalence of MLTCs as they are known to influence health [112]. It is possible that racism and multiple forms of discrimination can intersect with demographic factors (e.g. age, gender, and/or sexual orientation) resulting in disadvantage in accessing key economic, physical and social resources for some, thereby, leading to socioeconomic and health inequalities [113-115]. In turn, these inequalities can result in higher prevalence of some health conditions which may then accelerate the development of MLTCs in some minoritised ethnic group populations. Racism and multiple forms of discrimination can also lead to ethnic inequalities in healthcare access, utilisation and care quality [116] through a number of pathways. For example, findings from international studies suggest that negative discriminatory practices can result in mistrust of healthcare professionals, non-compliance with treatment, delayed diagnoses and treatment and even forgone healthcare [117-119]. These outcomes are detrimental as they not only exacerbate existing health inequalities, but they can also lead to the development and/or progression of MLTCs. However, these processes have received very little investigation in the context of ethnic inequalities in the prevalence of MLTCs in the UK.

### Strengths and limitations

In presenting the state of the evidence, this review has highlighted the inconsistent ways in which MLTCs are defined and measured with the number of long-term conditions varying between studies. This lack of consensus in how MLTCs are captured has been raised by authors of other reviews of prevalence of MLTCs in the general population [120, 121]. They argue that the operational definition of multimorbidity may impact prevalence estimates and call for the standardisation of the definition and assessment of multimorbidity so as to better understand the phenomenon [120, 121]. The review has also highlighted the limitations of the studies in this area. First, many did not provide age-adjusted prevalence estimates of MLTCs. Second, in many of these studies ethnic groups with small sample sizes and those with missing ethnicity data were often excluded from analyses. We, therefore, have an incomplete picture of ethnic inequalities across ethnic group populations in the UK. Relatedly, information on sensitivity analysis of missing ethnicity data was only available in four studies. Finally, in 18% of the studies, ethnicity was assigned by practitioners, clinicians, or researchers via a combination of name recognition software and genealogy. These modes of assigning ethnicity are problematic primarily because it may be incongruent with the ways in which individuals choose to identify themselves [122].

A limitation of this review is that a single reviewer initially screened the titles and abstracts, and excluded irrelevant studies given the large number of studies retrieved. Thereafter, a sample of the studies (10%) were double screened by BH and LB. As such, it is possible that some studies may have been inadvertently excluded. However, we manually searched the reference lists of key studies and relevant systematic reviews, thereby, reducing the likelihood of missing relevant studies. A strength of the review is that we conformed to the PRISMA guidelines to help transparently report the review process [20, 123]. We also conducted the electronic search across a range of databases to identify published and unpublished studies. Another strength of the review is that when synthesising the results of studies that contributed to the evidence of ethnic inequalities in the prevalence of MLTCs, we only included studies that adjusted for at least age because the risk of acquiring multiple health conditions increases with age [124]. In doing so, we reduced the likelihood of presenting results that are misleading.

## CONCLUSIONS

In this review, we have presented the state of the evidence on ethnic inequalities in the prevalence of MLTCs in the UK. We have identified and described the literature in this area and in doing so, illuminated the scope of work required to enhance future analyses of ethnic inequalities in people with MLTCs. With the exception of two studies that focused on diabetes, the studies identified point to the existence of ethnic inequalities in the prevalence of MLTCs. These studies also suggest that Indian, Pakistani, Bangladeshi, Black African, Black Caribbean, and people who identify as Black Other, Other Asian, and Mixed may be at higher risk of MLTCs. The majority of these studies used broad ethnic categories in their analyses. They also focused on different health conditions. Further, they drew on patient records which may inadvertently exclude those who are not in contact with healthcare services. Thus, the results provide a partial picture of ethnic inequalities in the prevalence of multiple conditions.

Given the complexity of multiple conditions, the diversity of the minoritised ethnic group populations in the UK, and the varied pathways through which they come to develop MLTCs, future studies would benefit from conceptualising and analysing the prevalence of ethnic inequalities through an intersectional lens. This work could shed light on the extent to which key explanatory pathways, including racism and discrimination, play a role in the development of MLTCs in different ethnic group populations. It is findings of analyses such as these that could help to inform strategies to reduce ethnic inequalities in the prevalence of MLTCs.

## Data Availability

All data produced in the present study are publicly available

## LIST OF ABBREVIATIONS

ASSIA: Applied Social Sciences Index and Abstracts
BAME: Black Asian and Minority Ethnic
BME: Black and Minority Ethnic
BMI: Body Mass Index
CI: Confidence Interval
COVID-19: Coronavirus Disease 2019
CPRD: Clinical Practice Research Datalink
EMBASE: Excerpta Medica data base
MLTCs: Multiple long-term conditions
NHS: National Health Service
OR: Odds Ratios
PRISMA: Preferred Reporting Items for Systematic Reviews and Meta-Analyses
THIN: The Health Improvement Network
UK: United Kingdom

## DECLARATIONS

### Ethics approval and consent to participate

Not Applicable

### Consent for publication

Not Applicable

### Availability of data and materials

The data extracted from the included studies are publicly available.

### Authors’ contributions

LB and MS devised the primary research goals and objectives of this systematic review. LB MS, and BH planned the methodological approach and developed the protocol. BH formulated the search terms together with LB and MS. BH performed the search, imported the identified studies and eliminated duplicate studies. BH and LB screened and extracted data from a random sample of studies (10%). BH screened and extracted data from the remaining studies (90%). BH narratively synthesised the findings with extensive methodological and intellectual feedback from LB and MS. BH prepared the manuscript and wrote the initial draft. LB and MS critically reviewed and commented on the initial and subsequent drafts. When reviewing the manuscripts, both LB and MS verified the data from the studies that contributed to the evidence of ethnic inequalities in the prevalence of multiple long□ term conditions. All authors had full access to the included studies. BH submitted the manuscript for publication. All authors have read and agreed to the published version of the manuscript.

## Acknowledgements

Not applicable

