## Supplementary files for "Ethnic inequalities in multiple long-term health conditions in the United Kingdom: a systematic review and narrative synthesis"

**SUPPLEMENTARY FILE 1: SEARCH TERMS**

**APPLIED SOCIAL SCIENCES INDEX AND ABSTRACTS**

| MLTCs + Ethnicity + Country | (#1 AND #2 AND #3) NOT #4 ((MAINSUBJECT.EXACT.EXPLODE("Mixed ethnicity") OR MAINSUBJECT.EXACT.EXPLODE("Ethnicity") OR ab,ti,if("Ethnic Group?" OR "african continental ancestry group" OR Arab OR Africa? OR Afro? OR Asian OR "Asian Continental Ancestry Group" OR “Asylum seeker” OR Bangladesh? OR Black OR “BME” OR “BAME” OR Caribbean OR China OR Chinese OR Cultur? OR Divers? OR Ethnic? OR Gypsy OR India? OR Irish OR Migrant OR Minorit? OR Mixed OR “Mixed ethnic?” OR “Multiple ethnic?” OR Multi$rac? OR ‘Other White’ OR Pakistan? OR Roma OR “White Other” OR Refugee? OR race OR racial? OR “South Asian” OR "European Continental Ancestry Group")) AND (MAINSUBJECT.EXACT("England and Wales") OR MAINSUBJECT.EXACT("Channel Islands") OR MAINSUBJECT.EXACT("UK") OR MAINSUBJECT.EXACT("Scotland") OR MAINSUBJECT.EXACT("England") OR MAINSUBJECT.EXACT("Northern Ireland") OR MAINSUBJECT.EXACT("Wales") OR ti,ab,if("United Kingdom" OR “UK” OR England OR Wales OR Scotland OR “Northern Ireland” OR Britain OR “Great Britain”)) AND (ti,ab,if(“Multiple Chronic Conditions” OR Co$morbid? OR Multi$morbidity OR Multi$patholog? OR “multiple condition?” OR “Multiple health condition?” OR “Multiple health problems” OR “Multiple medical conditions” OR “Multiple medical problems” OR “Pluri$patholog?” OR Polymorbid? OR “multiple illness?” OR “Multiple Chronic Health Conditions” or “Multiple Chronic Medical Conditions” OR “multiple chronic illness?”) OR MAINSUBJECT.EXACT.EXPLODE("Comorbidity"))) NOT (MAINSUBJECT.EXACT.EXPLODE("USA") OR MAINSUBJECT.EXACT.EXPLODE("North America") OR MAINSUBJECT.EXACT.EXPLODE("Canada") OR MAINSUBJECT.EXACT.EXPLODE("Australia") OR MAINSUBJECT.EXACT.EXPLODE("New Zealand") OR MAINSUBJECT.EXACT.EXPLODE("South America") OR MAINSUBJECT.EXACT.EXPLODE("Central America") OR ti,ab,if("Americas" OR "USA" OR America OR "North America" OR Canada OR Australia OR "New Zealand")) |
| --- | --- |
| excluded countries | MAINSUBJECT.EXACT.EXPLODE("USA") OR MAINSUBJECT.EXACT.EXPLODE("North America") OR MAINSUBJECT.EXACT.EXPLODE("Canada") OR MAINSUBJECT.EXACT.EXPLODE("Australia") OR MAINSUBJECT.EXACT.EXPLODE("New Zealand") OR MAINSUBJECT.EXACT.EXPLODE("South America") OR MAINSUBJECT.EXACT.EXPLODE("Central America") OR ti,ab,if("Americas" OR "USA" OR America OR "North America" OR Canada OR Australia OR "New Zealand") |
| Country | (MAINSUBJECT.EXACT("England and Wales") OR MAINSUBJECT.EXACT("Channel Islands") OR MAINSUBJECT.EXACT("UK") OR MAINSUBJECT.EXACT("Scotland") OR MAINSUBJECT.EXACT("England") OR MAINSUBJECT.EXACT("Northern Ireland") OR MAINSUBJECT.EXACT("Wales")) OR ti,ab,if("United Kingdom" OR “UK” OR England OR Wales OR Scotland OR “Northern Ireland” OR Britain OR “Great Britain”) |
| Ethnicity | (MAINSUBJECT.EXACT.EXPLODE("Mixed ethnicity") OR MAINSUBJECT.EXACT.EXPLODE("Ethnicity")) OR ab,ti,if("Ethnic Group?" OR "african continental ancestry group" OR Arab OR Africa? OR Afro? OR Asian OR "Asian Continental Ancestry Group" OR “Asylum seeker” OR Bangladesh? OR Black OR “BME” OR “BAME” OR Caribbean OR China OR Chinese OR Cultur? OR Divers? OR Ethnic? OR Gypsy OR India? OR Irish OR Migrant OR Minorit? OR Mixed OR “Mixed ethnic?” OR “Multiple ethnic?” OR Multi$rac? OR ‘Other White’ OR Pakistan? OR Roma OR “White Other” OR Refugee? OR race OR racial? OR “South Asian” OR "European Continental Ancestry Group") |
| MLTCs | ti,ab,if(“Multiple Chronic Conditions” OR Co$morbid? OR Multi$morbidity OR Multi$patholog? OR “multiple condition?” OR “Multiple health condition?” OR “Multiple health problems” OR “Multiple medical conditions” OR “Multiple medical problems” OR “Pluri$patholog?” OR Polymorbid? OR “multiple illness?” OR “Multiple Chronic Health Conditions” or “Multiple Chronic Medical Conditions” OR “multiple chronic illness?”) OR MAINSUBJECT.EXACT.EXPLODE("Comorbidity") |

**COCHRANE LIBRARY**

| MLTCs + Ethnicity + Country | (#1 AND #2 AND #3) NOT #4 |
| --- | --- |
| excluded countries | "Americas" OR “USA” OR America OR “North America” OR Canada OR Australia OR “New Zealand” |
| Country | "United Kingdom" OR “UK” OR England OR Wales OR Scotland OR “Northern Ireland” OR Britain OR “Great Britain” OR "channel Islands" |
| Ethnicity | "Ethnic Group*" OR "african continental ancestry group" OR Arab OR Africa* OR Afro* OR Asian OR "Asian Continental Ancestry Group" OR “Asylum seeker” OR Bangladesh* OR Black OR “BME” OR “BAME” OR Caribbean OR China OR Chinese OR Cultur* OR Divers* OR Ethnic* OR Gypsy OR India* OR Irish OR Migrant OR Minorit* OR Mixed OR “Mixed ethnic*” OR “Multiple ethnic*” OR Multirac* OR ‘Other White’ OR Pakistan? OR Roma OR “White Other” OR Refugee* OR race OR racial* OR “South Asian” OR "European Continental Ancestry Group" |
| MLTCs | “Multiple Chronic Conditions” OR Comorbid* OR Multimorbidity OR Multipatholog* OR “multiple condition*” OR “Multiple health condition*” OR “Multiple health problems” OR “Multiple medical conditions” OR “Multiple medical problems” OR “Pluripatholog*” OR Polymorbid* OR “multiple illness*” OR “Multiple Chronic Health Conditions” or “Multiple Chronic Medical Conditions” OR “multiple chronic illness*” |

**EMBASE, MEDLINE, PsycINFO**

| MLTCs + Ethnicity + Country | (#1 AND #2 AND #3) NOT #4 |
| --- | --- |
| Country | (United Kingdom) OR (UK) OR England OR Wales OR Scotland OR (Northern Ireland) OR Britain OR (Great Britain) OR (Channel Islands) OR (England adj2 Wales) |
| Ethnicity | (Ethnic Group*) OR (african continental ancestry group) OR Arab OR Africa* OR Afro* OR Asian OR (Asian Continental Ancestry Group) OR (Asylum seeker) OR Bangladesh* OR Black OR (BME) OR (BAME) OR Caribbean OR China OR Chinese OR Cultur* OR Divers* OR Ethnic* OR Gypsy OR India* OR Irish OR Migrant OR Minorit* OR Mixed OR (Mixed ethnic*) OR (Multiple ethnic*) OR Multirac* OR (Other White) OR Pakistan* OR Roma OR (White Other) OR Refugee* OR race OR racial* OR (South Asian) OR (European Continental Ancestry Group) |
| MLTCs | (Multiple Chronic Conditions) OR Comorbid* OR Multimorbidity OR Multipatholog* OR (multiple adj2 condition*) OR (Multiple health condition*) OR (Multiple health problem*) OR (Multiple medical condition*) OR (Multiple medical problems) OR Pluripatholog* OR Polymorbid* OR (multiple illness*) OR (Multiple Chronic Health Conditions) or (Multiple Chronic Medical Conditions) OR (multiple chronic illness*) OR (multiple adj2 problem) OR (multiple adj2 illness) |

**PUBMED**

| MLTCs + Ethnicity + Country | (#1 AND #2 AND #3) NOT #4 (("Multiple Chronic Conditions"[MeSH Terms] OR "Comorbidity"[MeSH Terms] OR "Multimorbidity"[MeSH Terms] OR "comorbid*"[Title/Abstract] OR "co morbid*"[Title/Abstract] OR "multimorbid*"[Title/Abstract] OR "multipatholog*"[Title/Abstract] OR "multi patholog*"[Title/Abstract] OR "multiple condition"[Title/Abstract] OR "Multiple health problems"[Title/Abstract] OR "Multiple medical conditions"[Title/Abstract] OR "Multiple medical problems"[Title/Abstract] OR "pluripatholog*"[Title/Abstract] OR "polymorbid*"[Title/Abstract] OR "multiple illness*"[Title/Abstract] OR "Multiple Chronic Health Conditions"[Title/Abstract] OR "Multiple Chronic Medical Conditions"[Title/Abstract] OR "multiple chronic illness*"[Title/Abstract]) AND ("Ethnic Groups"[MeSH Terms] OR "african continental ancestry group"[MeSH Terms] OR ("arabs"[MeSH Terms] OR "arabs"[Title/Abstract] OR "arab"[Title/Abstract]) OR "africa*"[Title/Abstract] OR "afro*"[Title/Abstract] OR ("Asian Continental Ancestry Group"[MeSH Terms] OR ("asian"[Title/Abstract] AND "continental"[Title/Abstract] AND "ancestry"[Title/Abstract] AND "group"[Title/Abstract]) OR "Asian Continental Ancestry Group"[Title/Abstract] OR "asian"[Title/Abstract] OR "asians"[Title/Abstract]) OR "Asian Continental Ancestry Group"[MeSH Terms] OR ("Refugees"[MeSH Terms] OR "Refugees"[Title/Abstract] OR ("asylum"[Title/Abstract] AND "seeker"[Title/Abstract]) OR "asylum seeker"[Title/Abstract]) OR ("benzoyl l arginine methyl ester"[Supplementary Concept] OR "BME"[Title/Abstract] OR "BAME"[Title/Abstract]) OR "bangladesh*"[Title/Abstract] OR ("african continental ancestry group"[MeSH Terms] OR ("african"[Title/Abstract] AND "continental"[Title/Abstract] AND "ancestry"[Title/Abstract] AND "group"[Title/Abstract]) OR "african continental ancestry group"[Title/Abstract] OR "black"[Title/Abstract] OR "african americans"[MeSH Terms] OR ("african"[Title/Abstract] AND "americans"[Title/Abstract]) OR "african americans"[Title/Abstract] OR "blacks"[Title/Abstract] OR "blackness"[Title/Abstract]) OR "BME"[Title/Abstract] OR "BAME"[Title/Abstract] OR ("caribbean s"[Title/Abstract] OR "caribbeans"[Title/Abstract] OR "west indies"[MeSH Terms] OR ("west"[Title/Abstract] AND "indies"[Title/Abstract]) OR "west indies"[Title/Abstract] OR "caribbean"[Title/Abstract] OR "caribbean region"[MeSH Terms] OR ("caribbean"[Title/Abstract] AND "region"[Title/Abstract]) OR "caribbean region"[Title/Abstract]) OR ("china"[MeSH Terms] OR "china"[Title/Abstract] OR "china s"[Title/Abstract] OR "chinas"[Title/Abstract]) OR ("Asian Continental Ancestry Group"[MeSH Terms] OR ("asian"[Title/Abstract] AND "continental"[Title/Abstract] AND "ancestry"[Title/Abstract] AND "group"[Title/Abstract]) OR "Asian Continental Ancestry Group"[Title/Abstract] OR "chinese"[Title/Abstract] OR "chineses"[Title/Abstract]) OR "cultur*"[Title/Abstract] OR "divers*"[Title/Abstract] OR ("Ethnic Groups"[MeSH Terms] OR ("ethnic"[Title/Abstract] AND "groups"[Title/Abstract]) OR "Ethnic Groups"[Title/Abstract] OR ("ethnic"[Title/Abstract] AND "group"[Title/Abstract]) OR "ethnic group"[Title/Abstract]) OR "ethnic*"[Title/Abstract] OR ("roma"[MeSH Terms] OR "roma"[Title/Abstract] OR "gypsies"[Title/Abstract] OR "gypsy"[Title/Abstract]) OR "india*"[Title/Abstract] OR "Irish"[Title/Abstract] OR ("migrant s"[Title/Abstract] OR "transients and migrants"[MeSH Terms] OR ("transients"[Title/Abstract] AND "migrants"[Title/Abstract]) OR "transients and migrants"[Title/Abstract] OR "migrant"[Title/Abstract] OR "migrants"[Title/Abstract]) OR "minorit*"[Title/Abstract] OR ("mixed"[Title/Abstract] OR "mixes"[Title/Abstract] OR "mixing"[Title/Abstract] OR "mixings"[Title/Abstract]) OR ("mixed"[Title/Abstract] OR "mixes"[Title/Abstract] OR "mixing"[Title/Abstract] OR "mixings"[Title/Abstract]) OR ("multiple"[Title/Abstract] OR "multiples"[Title/Abstract]) OR "multirac*"[Title/Abstract] OR "multi rac*"[Title/Abstract] OR ("other"[Title/Abstract] AND ("European Continental Ancestry Group"[MeSH Terms] OR ("european"[Title/Abstract] AND "continental"[Title/Abstract] AND "ancestry"[Title/Abstract] AND "group"[Title/Abstract]) OR "European Continental Ancestry Group"[Title/Abstract] OR "white"[Title/Abstract] OR "whites"[Title/Abstract])) OR "pakistan*"[Title/Abstract] OR ("roma"[MeSH Terms] OR "roma"[Title/Abstract]) OR (("European Continental Ancestry Group"[MeSH Terms] OR ("european"[Title/Abstract] AND "continental"[Title/Abstract] AND "ancestry"[Title/Abstract] AND "group"[Title/Abstract]) OR "European Continental Ancestry Group"[Title/Abstract] OR "white"[Title/Abstract] OR "whites"[Title/Abstract]) AND "other"[Title/Abstract]) OR "Refugees"[MeSH Terms] OR ("continental population groups"[MeSH Terms] OR ("continental"[Title/Abstract] AND "population"[Title/Abstract] AND "groups"[Title/Abstract]) OR "continental population groups"[Title/Abstract] OR "race"[Title/Abstract]) OR "racial*"[Title/Abstract] OR "South Asian"[Title/Abstract] OR "European Continental Ancestry Group"[MeSH Terms]) AND ("United Kingdom"[MeSH Terms] OR "UK"[Title/Abstract] OR "england"[MeSH Terms] OR "england"[Title/Abstract] OR "england s"[Title/Abstract] OR "englands"[Title/Abstract] OR "wales"[MeSH Terms] OR "wales"[Title/Abstract] OR "wales s"[Title/Abstract] OR "scotland"[MeSH Terms] OR "scotland"[Title/Abstract] OR "scotland s"[Title/Abstract] OR "Northern Ireland"[Title/Abstract] OR "britain"[Title/Abstract] OR "britain s"[Title/Abstract] OR "britains"[Title/Abstract] OR "Great Britain"[Title/Abstract])) NOT ("Americas"[MeSH Terms] OR "USA"[Title/Abstract] OR "America"[Title/Abstract] OR "North America"[Title/Abstract] OR "Canada"[Title/Abstract] OR "Australia"[Title/Abstract] OR "New Zealand"[Title/Abstract]) |
| --- | --- |
| excluded countries | "Americas"[Mesh] OR "USA" [Title/Abstract] OR America [Title/Abstract] OR "North America" [Title/Abstract] OR Canada [Title/Abstract] OR Australia [Title/Abstract] OR "New Zealand" [Title/Abstract] |
| Country | "United Kingdom"[MeSH Terms] OR "UK"[Title/Abstract] OR "england"[MeSH Terms] OR "england"[Title/Abstract] OR "england s"[Title/Abstract] OR "englands"[Title/Abstract] OR "wales"[MeSH Terms] OR "wales"[Title/Abstract] OR "wales s"[Title/Abstract] OR "scotland"[MeSH Terms] OR "scotland"[Title/Abstract] OR "scotland s"[Title/Abstract] OR "Northern Ireland"[Title/Abstract] OR "britain"[Title/Abstract] OR "britain s"[Title/Abstract] OR "britains"[Title/Abstract] OR "Great Britain"[Title/Abstract] |
| Ethnicity | "Ethnic Groups"[MeSH Terms] OR "african continental ancestry group"[MeSH Terms] OR ("arabs"[MeSH Terms] OR "arabs"[Title/Abstract] OR "arab"[Title/Abstract]) OR "africa*"[Title/Abstract] OR "afro*"[Title/Abstract] OR ("Asian Continental Ancestry Group"[MeSH Terms] OR ("asian"[Title/Abstract] AND "continental"[Title/Abstract] AND "ancestry"[Title/Abstract] AND "group"[Title/Abstract]) OR "Asian Continental Ancestry Group"[Title/Abstract] OR "asian"[Title/Abstract] OR "asians"[Title/Abstract]) OR "Asian Continental Ancestry Group"[MeSH Terms] OR ("Refugees"[MeSH Terms] OR "Refugees"[Title/Abstract] OR ("asylum"[Title/Abstract] AND "seeker"[Title/Abstract]) OR "asylum seeker"[Title/Abstract]) OR ("benzoyl l arginine methyl ester"[Supplementary Concept] OR "BME"[Title/Abstract] OR "BAME"[Title/Abstract]) OR "bangladesh*"[Title/Abstract] OR ("african continental ancestry group"[MeSH Terms] OR ("african"[Title/Abstract] AND "continental"[Title/Abstract] AND "ancestry"[Title/Abstract] AND "group"[Title/Abstract]) OR "african continental ancestry group"[Title/Abstract] OR "black"[Title/Abstract] OR "african americans"[MeSH Terms] OR ("african"[Title/Abstract] AND "americans"[Title/Abstract]) OR "african americans"[Title/Abstract] OR "blacks"[Title/Abstract] OR "blackness"[Title/Abstract]) OR "BME"[Title/Abstract] OR "BAME"[Title/Abstract] OR ("caribbean s"[Title/Abstract] OR "caribbeans"[Title/Abstract] OR "west indies"[MeSH Terms] OR ("west"[Title/Abstract] AND "indies"[Title/Abstract]) OR "west indies"[Title/Abstract] OR "caribbean"[Title/Abstract] OR "caribbean region"[MeSH Terms] OR ("caribbean"[Title/Abstract] AND "region"[Title/Abstract]) OR "caribbean region"[Title/Abstract]) OR ("china"[MeSH Terms] OR "china"[Title/Abstract] OR "china s"[Title/Abstract] OR "chinas"[Title/Abstract]) OR ("Asian Continental Ancestry Group"[MeSH Terms] OR ("asian"[Title/Abstract] AND "continental"[Title/Abstract] AND "ancestry"[Title/Abstract] AND "group"[Title/Abstract]) OR "Asian Continental Ancestry Group"[Title/Abstract] OR "chinese"[Title/Abstract] OR "chineses"[Title/Abstract]) OR "cultur*"[Title/Abstract] OR "divers*"[Title/Abstract] OR ("Ethnic Groups"[MeSH Terms] OR ("ethnic"[Title/Abstract] AND "groups"[Title/Abstract]) OR "Ethnic Groups"[Title/Abstract] OR ("ethnic"[Title/Abstract] AND "group"[Title/Abstract]) OR "ethnic group"[Title/Abstract]) OR "ethnic*"[Title/Abstract] OR ("roma"[MeSH Terms] OR "roma"[Title/Abstract] OR "gypsies"[Title/Abstract] OR "gypsy"[Title/Abstract]) OR "india*"[Title/Abstract] OR "Irish"[Title/Abstract] OR ("migrant s"[Title/Abstract] OR "transients and migrants"[MeSH Terms] OR ("transients"[Title/Abstract] AND "migrants"[Title/Abstract]) OR "transients and migrants"[Title/Abstract] OR "migrant"[Title/Abstract] OR "migrants"[Title/Abstract]) OR "minorit*"[Title/Abstract] OR ("mixed"[Title/Abstract] OR "mixes"[Title/Abstract] OR "mixing"[Title/Abstract] OR "mixings"[Title/Abstract]) OR ("mixed"[Title/Abstract] OR "mixes"[Title/Abstract] OR "mixing"[Title/Abstract] OR "mixings"[Title/Abstract]) OR ("multiple"[Title/Abstract] OR "multiples"[Title/Abstract]) OR "multirac*"[Title/Abstract] OR "multi rac*"[Title/Abstract] OR ("other"[Title/Abstract] AND ("European Continental Ancestry Group"[MeSH Terms] OR ("european"[Title/Abstract] AND "continental"[Title/Abstract] AND "ancestry"[Title/Abstract] AND "group"[Title/Abstract]) OR "European Continental Ancestry Group"[Title/Abstract] OR "white"[Title/Abstract] OR "whites"[Title/Abstract])) OR "pakistan*"[Title/Abstract] OR ("roma"[MeSH Terms] OR "roma"[Title/Abstract]) OR (("European Continental Ancestry Group"[MeSH Terms] OR ("european"[Title/Abstract] AND "continental"[Title/Abstract] AND "ancestry"[Title/Abstract] AND "group"[Title/Abstract]) OR "European Continental Ancestry Group"[Title/Abstract] OR "white"[Title/Abstract] OR "whites"[Title/Abstract]) AND "other"[Title/Abstract]) OR "Refugees"[MeSH Terms] OR ("continental population groups"[MeSH Terms] OR ("continental"[Title/Abstract] AND "population"[Title/Abstract] AND "groups"[Title/Abstract]) OR "continental population groups"[Title/Abstract] OR "race"[Title/Abstract]) OR "racial*"[Title/Abstract] OR "South Asian"[Title/Abstract] OR "European Continental Ancestry Group"[MeSH Terms] |
| MLTCs | "Multiple Chronic Conditions"[MeSH Terms] OR "Comorbidity"[MeSH Terms] OR "Multimorbidity"[MeSH Terms] OR "comorbid*"[Title/Abstract] OR "co morbid*"[Title/Abstract] OR "multimorbid*"[Title/Abstract] OR "multipatholog*"[Title/Abstract] OR "multi patholog*"[Title/Abstract] OR "multiple condition"[Title/Abstract] OR "Multiple health condition"[Title/Abstract] OR "Multiple health problems"[Title/Abstract] OR "Multiple medical conditions"[Title/Abstract] OR "Multiple medical problems"[Title/Abstract] OR "pluripatholog*"[Title/Abstract] OR "polymorbid*"[Title/Abstract] OR "multiple illness*"[Title/Abstract] OR "Multiple Chronic Health Conditions"[Title/Abstract] OR "Multiple Chronic Medical Conditions"[Title/Abstract] OR "multiple chronic illness*"[Title/Abstract] |

**SCIENCE DIRECT**

| Multimorbidity + Ethnicity + Country | (Comorbid OR Multimorbid OR "multiple health conditions" OR polymorbid) AND (ethnic OR race ) AND ("United kingdom" OR "Great Britain") |
| --- | --- |

**SCOPUS**

| MLTCs + Ethnicity + Country | (#1 AND #2 AND #3) NOT #4 ( ( TITLE-ABS-KEY ( ( "multiple chronic conditions" ) OR comorbid* OR multimorbidity OR multipatholog* OR ( "multiple condition*" ) OR ( "multiple health condition*" ) OR ( "multiple health problem*" ) OR ( "multiple medical condition*" ) OR ( "multiple medical problems" ) OR pluripatholog* OR polymorbid* OR ( "multiple W/1 illness*" ) OR ( "multiple chronic health conditions" ) OR ( "multiple chronic medical conditions" ) OR ( "multiple chronic illness*" ) OR ( "multiple PRE/2 problem" ) OR ( "multiple PRE/2 illness" ) ) ) AND ( TITLE-ABS-KEY ( ( "ethnic group*" ) OR ( "african continental ancestry group" ) OR arab OR africa* OR afro* OR asian OR ( "asian continental ancestry group" ) OR ( "asylum seeker" ) OR bangladesh* OR black OR ( bme ) OR ( bame ) OR caribbean OR china OR chinese OR cultur* OR divers* OR ethnic* OR gypsy OR india* OR irish OR migrant OR minorit* OR mixed OR ( "mixed ethnic*" ) OR ( "multiple ethnic*" ) OR multirac* OR ( "other white" ) OR pakistan* OR roma OR ( white W/1 other ) OR refugee* OR race OR racial* OR ( "south Asian" ) OR ( "european continental ancestry group" ) ) ) AND ( TITLE-ABS-KEY ( ( "united kingdom" ) OR ( uk ) OR england OR wales OR scotland OR ( "northern Ireland" ) OR britain OR ( "great Britain" ) OR ( "channel islands" ) OR ( "england and wales" ) ) ) ) AND NOT ( TITLE-ABS-KEY ( americas OR ( "USA" ) OR america OR ( "North America" ) OR canada OR australia OR ( "New Zealand" ) ) ) |
| --- | --- |
| excluded countries | TITLE-ABS-KEY (Americas OR ("USA") OR America OR ("North America”) OR Canada OR Australia OR ("New Zealand")) |
| Country | TITLE-ABS-KEY ( ( "united kingdom" ) OR ( uk ) OR england OR wales OR scotland OR ( "northern Ireland" ) OR britain OR ( "great Britain" ) OR ( "channel islands" ) OR ( "england and wales" ) ) |
| Ethnicity | TITLE-ABS-KEY ( ( "ethnic group*" ) OR ( "african continental ancestry group" ) OR arab OR africa* OR afro* OR asian OR ( "asian continental ancestry group" ) OR ( "asylum seeker" ) OR bangladesh* OR black OR ( bme ) OR ( bame ) OR caribbean OR china OR chinese OR cultur* OR divers* OR ethnic* OR gypsy OR india* OR irish OR migrant OR minorit* OR mixed OR ( "mixed ethnic*" ) OR ( "multiple ethnic*" ) OR multirac* OR ( "other white" ) OR pakistan* OR roma OR ( white W/1 other ) OR refugee* OR race OR racial* OR ( "south Asian" ) OR ( "european continental ancestry group" ) ) |
| MLTCs | TITLE-ABS-KEY ( ( "multiple chronic conditions" ) OR comorbid* OR multimorbidity OR multipatholog* OR ( "multiple condition*" ) OR ( "multiple health condition*" ) OR ( "multiple health problem*" ) OR ( "multiple medical condition*" ) OR ( "multiple medical problems" ) OR pluripatholog* OR polymorbid* OR ( "multiple W/1 illness*" ) OR ( "multiple chronic health conditions" ) OR ( "multiple chronic medical conditions" ) OR ( "multiple chronic illness*" ) OR ( "multiple PRE/2 problem" ) OR ( "multiple PRE/2 illness" ) ) |

**WEB OF SCIENCE CORE COLLECTION**

| MLTCs + Ethnicity + Country | (#1 AND #2 AND #3) |
| --- | --- |
| Country | TOPIC: (( "united kingdom" ) OR ( uk )  OR England OR wales OR scotland OR ( "northern Ireland" ) OR britain  OR ( "great Britain" ) OR ( "channel islands" ) OR ( "england and wales" ) |
| Ethnicity | TOPIC: ((Ethnic Group*) OR (african continental ancestry group)  OR Arab OR Africa* OR Afro* OR Asian OR (Asian Continental ancestry Group) OR (Asylum seeker) OR Bangladesh* OR Black OR (BME) OR (BAME) OR Caribbean OR China OR Chinese OR Cultur* OR  Divers* OR Ethnic* OR Gypsy OR India* OR Irish OR Migrant OR Minorit* OR Mixed OR (Mixed ethnic*) OR (Multiple ethnic*) OR Multirac* OR (Other White) OR Pakistan* OR Roma OR (White Other)  OR Refugee* OR race OR racial* OR South Asian) OR (European Continental Ancestry Group) ) |
| MLTCs | TOPIC:((Multiple Chronic Conditions) OR Comorbid* OR Multimorbidity OR Multipatholog* OR (multiple condition*) OR (Multiple health condition*) OR (Multiple health problems) OR (Multiple medical conditions) OR (Multiple medical problems) OR  (Pluripatholog*) OR Polymorbid* OR (multiple illness*) OR (Multiple Chronic Health Conditions) OR (Multiple Chronic Medical Conditions)  OR (multiple chronic illness*) ) |

**OPENGREY**

| MLTCs + Ethnicity + Country | (Comorbid OR Multimorbid OR "multiple health conditions" OR polymorbid) AND (ethnic OR race ) AND ("United kingdom" OR "Great Britain") |
| --- | --- |

**SUPPLEMENTARY FILE 2**

**Figure showing the number of studies by the number of ethnic group categories**

Number of Studies

| Number of ethnic group categories | Field2 |
| --- | --- |
| 2 ethnic group categories | 18 |
| 3 ethnic group categories | 24 |
| 4 ethnic group categories | 18 |
| 5 ethnic group categories | 13 |
| 6 ethnic group categories | 7 |
| 7 ethnic group categories | 2 |
| 8 ethnic group categories | 1 |
| 9 ethnic group categories | 1 |
